# Estimation of monkeypox spread in a non-endemic country considering contact tracing and self-reporting: a stochastic modeling study

**DOI:** 10.1101/2022.08.11.22278654

**Authors:** Youngsuk Ko, Renier Mendoza, Victoria May Mendoza, Yubin Seo, Jacob Lee, Eunok Jung

## Abstract

**Background:** In May 2022, monkeypox started to spread in non-endemic countries. After the number of confirmed cases reached more than 16,000 in July, the World Health Organization declared the highest alert over the outbreak.

**Methods:** To investigate the effects of contact tracing and self-reporting of primary cases in the local community, a stochastic model is developed. A delay simulation algorithm based on Gillespie’s stochastic chemical kinetics is used to quantify the number of infections, contacts made by the infectors, and duration from the arrival of the primary case until the detection of the index case (and until there are no more local infections), under various scenarios.

**Results:** We found that if the primary case does not self-report, taking into account a population of 10,000, the average number of infections could range from 30 to 67, while the number of contacts made by infectors could range from 221 to 498. On the other hand, if the primary case self-reports, the average number of infections and contacts could range from 5 to 7 and 40 to 52, respectively. The average duration from the primary case arrival until the first index case detection (or until there are no more local infections) ranged from 8 to 10 days (18 to 21 days) if the primary case does not self-report, and approximately 3 days (8 days) if the primary case self-reports. Moreover, if the number of close contacts per day is doubled in our simulation settings, then the number of infections could increase by 53%.

**Conclusion:** The number and duration of the infections are strongly affected by the self-reporting behavior of the primary case and the delay in the detection of the index case. Our study emphasizes the importance of border control, which aims to immediately detect the primary case before secondary infections occur.

## Introduction

Monkeypox is a zoonotic disease caused by the monkeypox virus. Two possible means of transmission are animal-to-human and human-to-human. Animal-to-human transmission, also known as zoonotic transmission, is possible upon contact with or consumption of an infected natural host animal, such as rats, squirrels, and prairie dogs.^1,2^ Human-to-human transmission can be caused by respiratory droplets or contact with lesions and bodily fluids. Contact with contaminated materials such as clothing, beddings, or eating utensils can also cause infections.^3^ The signs and symptoms of monkeypox are similar to smallpox but are less severe. In the current outbreak, symptoms are described as flu-like, followed by a rash starting in the genital and perianal areas, which may or may not spread to the other parts of the body.^4,5^

The monkeypox virus was first reported in 1970.^6^ It is commonly found in Central and West Africa and occasionally identified in other countries.^1^ If undetected, transmission in non-endemic countries is possible, triggering outbreaks.^7,8^ Since May 2022, an unprecedented outbreak of human monkeypox cases has been observed in Australia and several countries in Europe and North America.^9^ On July 23, World Health Organization declared a global health emergency as the number of confirmed cases reached more than 16,000.^10^ As of August 11, 2022, Unted States recorded the highest number of cumulative cases (10,360), followed by Spain (5,482) and Germany (3,025).^11^

The basic reproductive number of monkeypox was estimated to be 2.13 (uncertainty bounds 1.46–2.67).^12^ Although it is less than the basic reproductive number of COVID-19, monkeypox has a potential to become an epidemic.^13^ Hence, understanding the transmission dynamics of the disease is essential to mitigate its spread. There are only a few works on monkeypox transmission models.^14,15,16,17,18^ These models include both types of transmissions in endemic regions. However, since the disease had spread in non-endemic countries where natural animal hosts are not present, these models may not be applicable.

The social media coverage about the monkeypox spread directly or indirectly generates racist and homophobic stereotypes that worsen stigma.^9^ Stigma can discourage people from seeking medical help, which may impede the efforts in identifying cases.^19,20^ Self-reporting plays an important role in mitigating the spread of infections. Once a suspected case is identified, the WHO recommends that contact identification and contact tracing be initiated.^21^ In this study, we are interested in understanding the dynamics of monkeypox infection in non-endemic settings. Using a stochastic model, we would like to discuss the following relevant questions:

1. Upon the arrival of a primary case in a non-endemic country, how many infections may occur?^22^
2. Depending on when the index case is found, how many contacts would the infector have already made?
3. How long is the duration of the outbreak?
4. What happens if the primary case does not self-report?

In this study, we estimated the potential monkeypox outbreak in a non-endemic country using a stochastic modeling method. Contact tracing, which is a well-known nonpharmaceutical intervention, and the behavior of the primary case, whether self-reporting or not, are considered in our simulation study.

## Methods

A stochastic model that assumes a nondelayed Markovian frequency-dependent disease transmission and incorporates delayed non-Markovian reactions is developed. The flow diagram of the model is displayed in Figure 1. The transmission rate consists of the number of contacts per unit time (*c*) and probability of successful disease transmission (*p*).^23^ Contact is made between infectors (*I*_1_ or *I*_2_) and susceptible hosts (*S*), formulated as the term 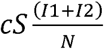, where *N* indicates the total population except isolated hosts (*S + C + E + I*_1_ + *I*_2_ *+ R*) and subscripts 1 or 2 denote self-reporting or non-self-reporting, respectively. Depending on *p*, a susceptible host who had contact with an infector can either be uninfected (*C*) or in latent stage (*E*). The probability that an infectee will later self-report is denoted by *ρ*.

**Figure 1.**
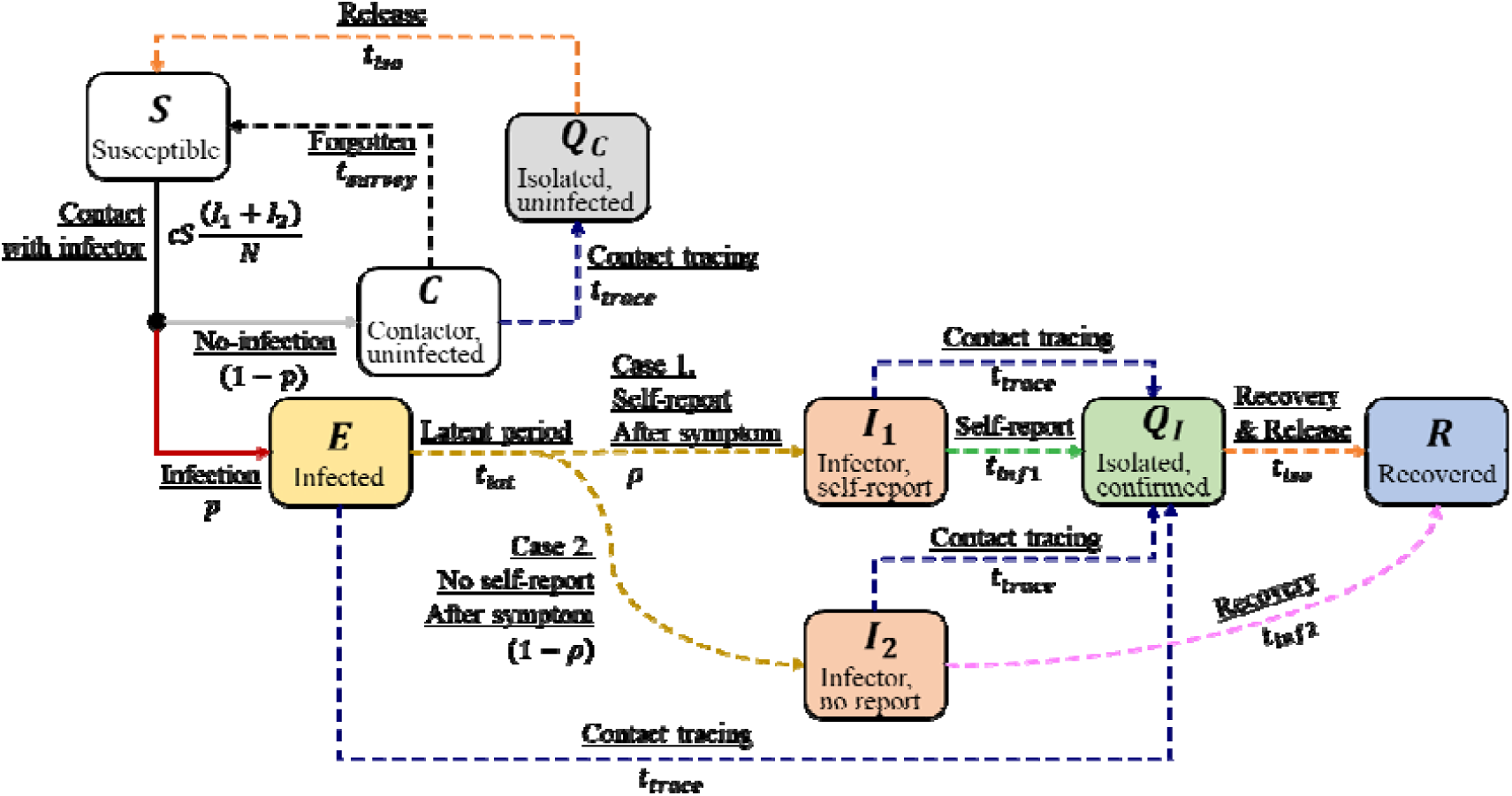
Flow diagram of the stochastic modeling. Note that arrows with solid lines indicate nondelayed (Markovian) reactions while arrows with dashed curves are delayed (non-Markovian) reactions.

When an infectee in the latent stage becomes an infector (*I*_1_ or *I*_2_), the host can spread the disease. If the infector self-reports (*I*_1_), then the host is isolated (*Q*_*I*_) and contact tracing events are generated once the case is confirmed. A certain proportion (*e*_*c*_) of hosts (*C, E, I*_1_, *I*_2_) who had contact with the confirmed host within contact survey time (*t*_*survey*_, 14 days, assumed) will be traced. An infector in *I*_2_ will stay in *I*_2_ until the host is naturally recovered (*R*), or until tracked and isolated. Note that contact tracing events are also generated if a traced host is infected (*E, I*_1_, *I*_2_). A host in *C* can be traced and isolated (*Q*_*C*_) even if the host is not infected or go back so S after *t*_*survey*_. We assumed the isolation time *t*_*iso*_ is 21 days.

Aggregating the values from Prem’s study, the average number of contacts per day (*c*) in Korea is estimated as 12.74 (3.20 household contacts, 9.54 else).^24^ Combining these contacts with the probability of successful disease transmission through a household contact and other contacts, 25.25% and 9.56%, respectively, the adjusted probability of successful disease transmission through a contact (*p*) is estimated as 13.51%.^25^ Denoting by *c*_*hh*_ and *c*_*other*_ (*p*_*hh*_ and *p*_*other*_) the daily number of (probability of infection through) household and other contacts, respectively, the probability of infection through a contact is calculated as follows,

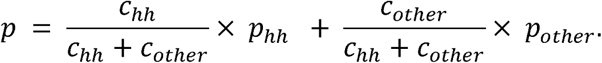

In our model, contact (resulting to non-infection or infection) is the only nondelayed reaction (Markovian, solid arrow line in Figure 1). The other reactions are delayed (Non-Markovian, dashed arrow). We set uniform distributions for delays that are uncertain but ranged (infectious period of non-self-reporting infector, *t*_*inf*2_, 14 to 28 days) or assumed (contact tracing time, *t*_*trace*_, 1 to 3 days).^26,27^ Latent period (*t*_*lat*_) and self-report time (*t*_*inf*1_) are generated from log-normal distribution and aggregated from previous studies.^28,29^ The average values of the latent period and self-report time are 8.5 and 2.96 days, respectively. We assumed that the delay for a host with monkeypox to self-report (*t*_*inf*1_) is similar to the period from symptom onset to diagnosis of a known case of COVID-19 exposure. The delays *t*_*survey*_ and *t*_*iso*_ are fixed to constant values.

Here, the definition of the primary case and index case are as follows. ^22^

✓ Primary case: the person who brings the disease to a community
✓ Index case: the first patient identified by health authorities with the disease

Considering whether the primary case self-reports or not (initial condition of the simulation is *I*_1_ or *I*_2_), self-report rate of infectees in the local community (*ρ*, 0.5 or 1), and contact tracing coverage (*e*_*c*_, 0.5 or 1), we set up eight scenarios, ran 10,000 simulations for each scenario, and observed the number of infections, contacts, duration of primary case arrival to index case detection (***P1***), and duration of primary case arrival to the end of the simulation (***P2***). End of the simulation means that all who were infected in the local community (except isolated) are removed, that is, *E + I*_1_ *+ I*_2_ = 0. The exact stochastic simulation algorithm for systems with delays was adopted to express the delayed events. A detailed description of the algorithm, which was developed based on Gillespie’s stochastic chemical kinetics, is presented in ^30^. We simply set the population size as 10,000.

## Results

The different scenarios are labeled ***S-1*** to ***S-8***, according to the setting of the simulation. The setting of each scenario and corresponding simulation results, illustrated as boxplots, are shown in Figure 2. Note that ***P1*** and ***P2*** denote the duration of the primary case arrival to index case detection and end of the simulation, respectively. Numerical results are summarized in Table 1.

**Table 1.** Simulation results of scenarios.

| Time phase | What a primary case do | Self-report rate of infectee | Contact trace coverage | Number of infection |  |  | Number of contact |  |  | Time duration (day) |  |  |
| --- | --- | --- | --- | --- | --- | --- | --- | --- | --- | --- | --- | --- |
|  |  |  |  | Mean | Median | 95% C.I. | Mean | Median | 95% C.I. | Mean | Median | 95% C.I. |
| From primary case arrival to index case | Self-report | 100% | 100% | 4.97 | 3 | [0,19] | 36.65 | 26 | [4,134] | 2.67 | 2 | [0.37,8.16] |
|  |  |  | 50% | 5.04 | 3 | [0,20] | 37.15 | 26 | [3,140] | 2.68 | 2 | [0.36,8.32] |
|  |  | 50% | 100% | 5.43 | 3 | [0,25] | 40.35 | 26 | [3,178] | 2.77 | 2 | [0.37,9.06] |
|  |  |  | 50% | 5.4 | 3 | [0,24] | 39.88 | 26 | [4,175] | 2.74 | 2 | [0.36,8.98] |
|  | No self-report | 100% | 100% | 19.76 | 18 | [8,40] | 146.23 | 136 | [68,283] | 8.22 | 8 | [4.68,12.19] |
|  |  |  | 50% | 19.59 | 18 | [8,39] | 145.31 | 136 | [67,275] | 8.2 | 8 | [4.71,12.23] |
|  |  | 50% | 100% | 30.97 | 26 | [9,83] | 229.2 | 191 | [77,602] | 9.55 | 9 | [5.17,14.67] |
|  |  |  | 50% | 30.94 | 26 | [9,81] | 228.46 | 190 | [77,590] | 9.55 | 9 | [5.23,14.7] |
| From primary case arrival to situation end | Self-report | 100% | 100% | 5.39 | 3 | [0,25] | 39.77 | 26 | [4,173] | 7.79 | 8 | [0.45,16.08] |
|  |  |  | 50% | 5.6 | 3 | [0,27] | 41.32 | 26 | [3,191] | 7.81 | 8 | [0.45,16.74] |
|  |  | 50% | 100% | 6.69 | 3 | [0,40] | 49.62 | 26 | [3,286] | 8.06 | 8 | [0.45,19.23] |
|  |  |  | 50% | 7.03 | 3 | [0,46] | 51.89 | 26 | [4,323] | 8.07 | 8 | [0.44,19.88] |
|  | No self-report | 100% | 100% | 29.86 | 27 | [12,62] | 221.2 | 201 | [105,447] | 17.56 | 18 | [12.7,22.76] |
|  |  |  | 50% | 31.61 | 29 | [12,66] | 234.65 | 214 | [106,476] | 17.84 | 18 | [12.6,23.36] |
|  |  | 50% | 100% | 57.48 | 45 | [14,172] | 425.49 | 332 | [118,1260] | 20.07 | 20 | [13.4,27.39] |
|  |  |  | 50% | 67.39 | 52 | [15,209] | 498.01 | 385 | [119,1533] | 20.92 | 21 | [13.56,28.68] |

**Figure 2.**
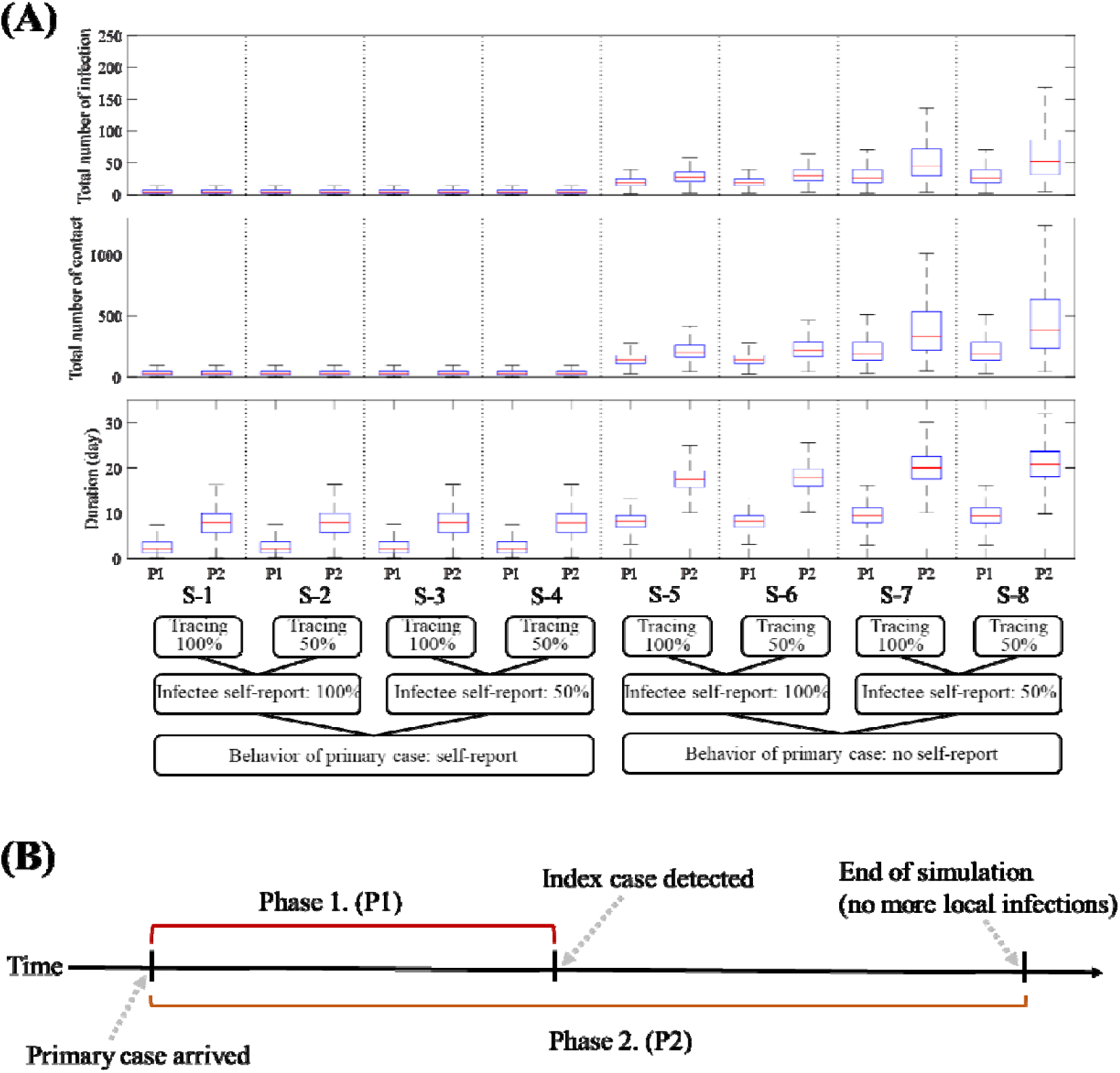
Scenario-based simulation results. (A) Boxplots showing the total number of infections, the total number of contacts, and duration of ***P1*** and ***P2*** corresponding to each scenario. (B) Timeline illustrating the two phases ***P1*** and ***P2***.

We observe that if the primary case does not self-report (***S-5*** to ***S-8***), then the simulations are worse than in the scenarios where the primary case self-reports (***S-1*** to ***S-4***). The average number of infections, contacts, and duration from primary case arrival to the end of simulation for scenarios ***S-1*** to ***S-4*** are 6.22, 45.99, and 7.94 days, respectively, which are 7.49, 7.50, and 2.4 times higher than for scenarios ***S-5*** to ***S-8*** (46.58, 344.75, and 19.10 days, respectively).

Considering the number of contacts per day and the probability of successful disease transmission, the expected number of infections per day is 1.72 (Number of contacts per day × probability of infection by contact). This means that in scenarios ***S-5*** to ***S-8***, where the primary case is not the index case, the primary case would already have made dozens of secondary infections until the index case is detected, which takes (incubation period) + (self-report time) days. The average duration of ***P1*** in scenarios ***S-1*** to ***S-4*** is 1 to 2 days since the primary case is most likely identical to the index case, while in ***S-5*** to ***S-8***, it takes 7 to 9 days on average until index case is detected.

Figure 3 shows the probability density of the total number of infections and contacts, and durations of ***P1*** and ***P2*** for scenarios ***S-3, S-4***, and ***S-5***. Although ***S-5*** is set to have 100% coverage for contact tracing and self-reporting rate of infectees, it shows the worst outcomes among the three scenarios. We observe skewed distributions for ***S-3*** and ***S-4***, suggesting that the situation would most likely be small scale. The average number of infections until the end of the simulation in ***S-5*** (29.86) is approximately more than four times higher than in ***S-3*** or ***S-4*** (6.69 or 7.03, respectively). Scenario ***S-4*** shows a slightly higher average number of infections compared to ***S-3*** due to the reduced contact tracing coverage.

**Figure 3.**
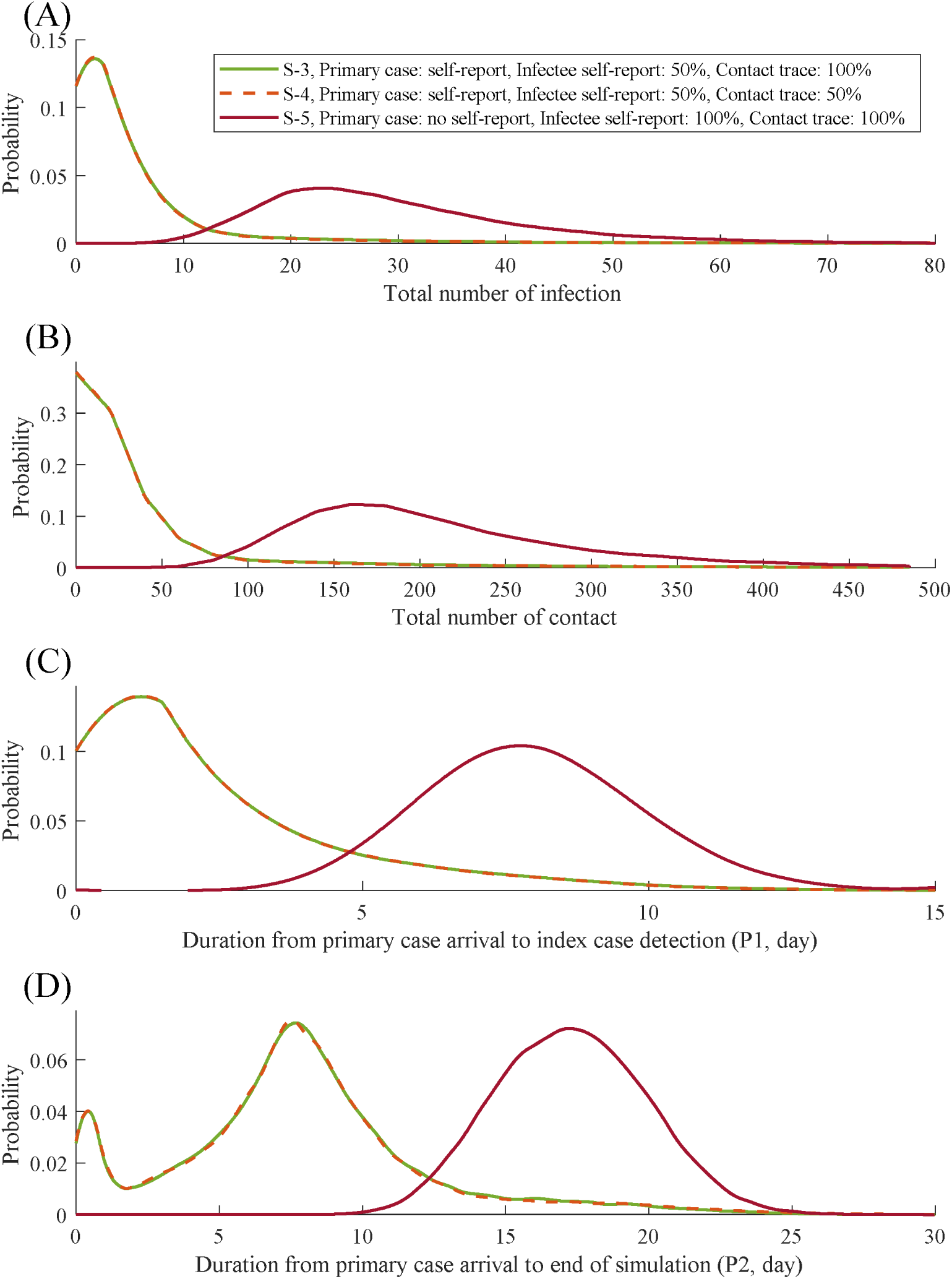
Distribution of the simulation results of three scenarios. (A) Total number of infections. (B) Total number of contacts. (C) Duration from primary case arrival to the identification of index case. (D) Duration from primary case arrival to end of the simulation.

To investigate what happens if the average number of household contacts is higher, such as in a potential super-spreader event, we considered doubling the number of household contacts for scenarios S-1 and S-5, which are denoted by ***S-1x*** and ***S-2x***. The plots in Figure 4 show the simulation results. As the number of household contacts doubled from 3.20 to 6.41, the average number of contacts and probability of disease transmission by contact increased to 15.94% and 15.87% (from 12.74 and 13.51%), respectively. The average number of infections and contacts of S-1x (S-5x) increased by 53.11% (46.67%) and 30.41% (24.72%), respectively. There were no significant changes in the durations of ***P1*** and ***P2***.

**Figure 4.**
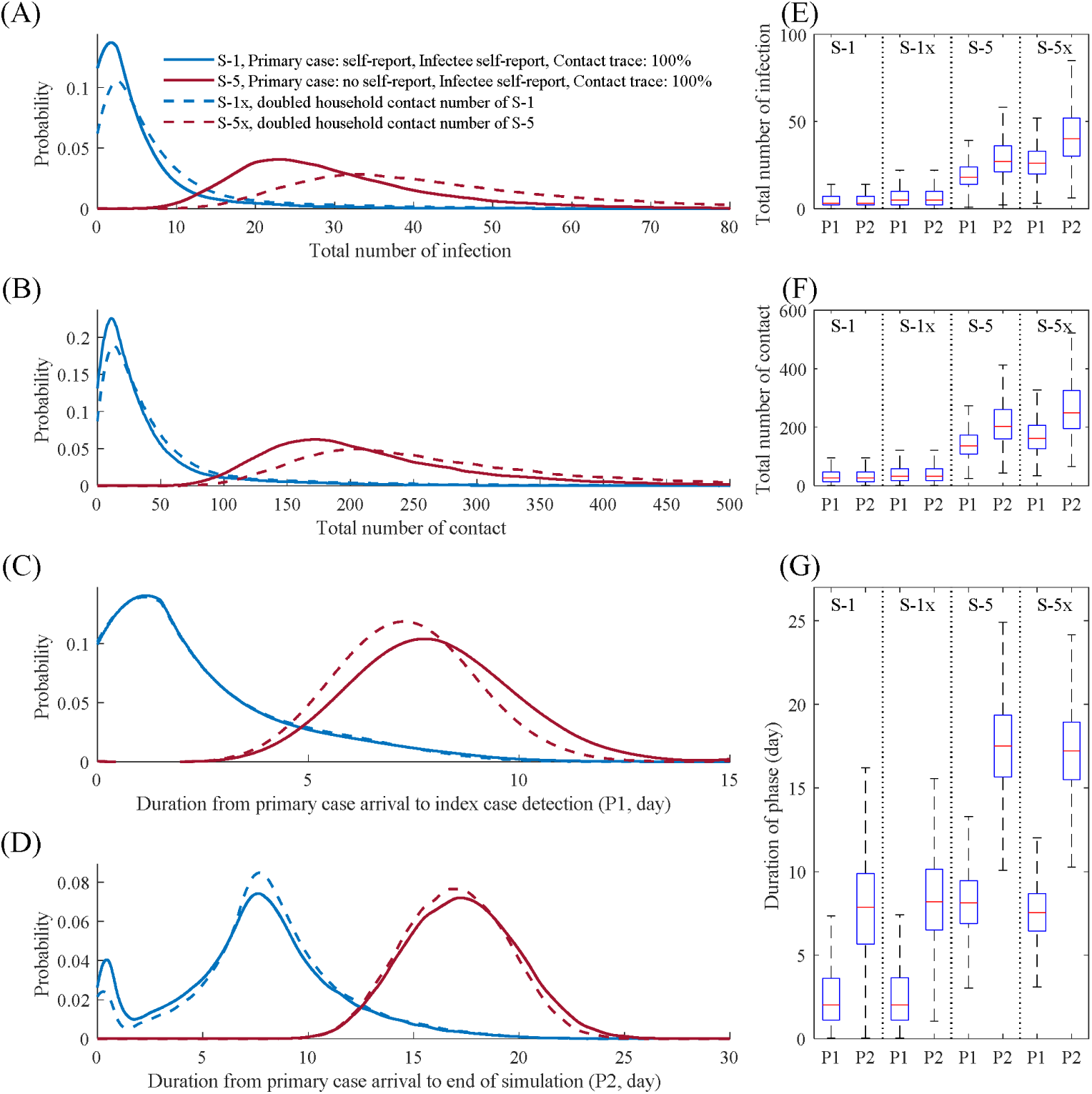
Distribution profile of simulation results if the number of household contacts is doubled. (A) Total number of infections. (B) Total number of contacts. (C) Duration from primary case arrival to the identification of the index case. (D) Duration from primary case arrival to end of the simulation. (E) Boxplot of (A), including the phase from primary case arrival to the identification of the index case. (F) Boxplot of (B), including the phase from primary case arrival to the identification of the index case. (G) Boxplot of (C) and (D).

## Discussion

In Korea, the first monkeypox case had self-reported right after arrival.^31^ As a result, there were no recorded secondary infections, which would be similar to a ‘good’ scenario (***S-1*** to ***S-4***) in our simulation setting. However, if the primary case was in the incubation stage and did not self-report after symptom onset, then hundreds of contacts and dozens of infections could be expected to have occurred in the local community once the index case was found. In this scenario, more efforts would have been needed to control the disease.

We investigated the effects of doubling the number of household contacts per day (which can be referred to as close contact). The results illustrated in Figure 4 warn of the potential risk of a significant increase in the number of monkeypox infections when unreported initial cases contacted more people, or if the group the primary case belongs to has more close contacts.

### Strengths and limitations

Our simulation results may guide the healthcare authorities in the planning and management of possible monkeypox outbreaks in non-endemic countries. For instance, it was shown that the delay in the detection of an index case is directly related to the number of contacts made, and knowing the scale of contact tracing efforts to be performed is crucial in managing manpower once the index case is found. In the simulations, we have used the average number of contacts per day suited for Korea but this can easily be adapted to other countries’ contact patterns. If the average household size is bigger or the number of contacts is more, then the probability of disease transmission increases.

There were limitations to our study. Contact tracing was applied to the model through a randomized sampling and not based on an individual’s network. Also, we assumed that contact tracing is perfect, that is, all infectors are found if 100% coverage is in effect. The probability of successful infection after contact might be overestimated since the concept of contact from the cited literature might differ from the definition of contact used in this study. Finally, we did not consider sexual contact in this study due to the limit of data. These limitations will be considered in the next study.

## Conclusions

In this study, the potential risk of monkeypox spread in a non-endemic country was studied using a stochastic model. We simulated outbreaks caused by a primary case that either self-reports or not and considered different coverage rates of contact tracing and self-reporting rates of infectees.

Stigma, which may result in an increase in hidden cases and negatively impact disease control, can be avoided by implementing evidence-based and rights-based approaches.^19^ Scientists have emphasized that knowledge about the epidemiology of the disease and awareness of its risks are important ways to prevent transmission and.^7,20^ Therefore, to encourage self-reporting, healthcare authorities must ensure confidentiality of confirmed cases and individuals under investigation, and access to health services. Moreover, prompt case finding and information campaigns must be conducted.

Our simulation results strongly emphasize the importance of border control to find the primary case. If the primary case self-reports, then despite having a low self-reporting rate of infectees and contact tracing rate, the situation does not become worse compared to the scenarios where the primary case does not self-report (in Figure 3).

## Data Availability

No new data were generated or analyzed in support of this research.

## Authors’ contributions

EJ acquired funding for this study. YK, JL, and EJ conceived and designed the study. YK, RM, and VM analyzed the data and wrote the first draft. EJ, JL, and YS gave suggestions on improving the quality of the analysis and monitored the study progress. All authors contributed to the data collection, checking and processing. All authors reviewed the final version of the manuscript. All authors read and approved the final manuscript.

